# Probabilistic modelling of effects of antibiotics and calendar time on transmission of healthcare-associated infection

**DOI:** 10.1101/2020.04.10.20060384

**Authors:** Mirjam Laager, Ben S Cooper, David W Eyre, on behalf of the CDC Modeling Infectious Diseases in Healthcare Program (MInD-Healthcare)

**Affiliations:** Nuffield Department of Medicine, University of Oxford, UK; Big Data Institute, Nuffield Department of Population Health, University of Oxford, UK

## Abstract

Healthcare-associated infection and antimicrobial resistance are major concerns. However, the extent to which antibiotic exposure affects transmission and detection of infections such as MRSA is unclear. Additionally, temporal trends are typically reported in terms of changes in incidence, rather than analysing underling transmission processes. We present a data-augmented Markov chain Monte Carlo approach for inferring changing transmission parameters over time, screening test sensitivity, and the effect of antibiotics on detection and transmission. We expand a basic model to allow use of typing information when inferring sources of infections. Using simulated data, we show that the algorithms are accurate, well-calibrated and able to identify antibiotic effects in sufficiently large datasets. We apply the models to study MRSA transmission in an intensive care unit in Oxford, UK with 7924 admissions over 10 years. We find that falls in MRSA incidence over time were associated with decreases in both the number of patients admitted to the ICU colonised with MRSA and in transmission rates. In our inference model, the data were not informative about the effect of antibiotics on risk of transmission or acquisition of MRSA, a consequence of the limited number of possible transmission events in the data. Our approach has potential to be applied to a range of healthcare-associated infections and settings and could be applied to study the impact of other potential risk factors for transmission. Evidence generated could be used to direct infection control interventions.

## Introduction

There is widespread concern that the rise of antimicrobial resistance (AMR) threatens the delivery of safe healthcare. This is particularly disconcerting as many of the causes of healthcare associated infection are themselves antimicrobial resistant. For example, methicillin-resistant *Staphylococcus aureus* (MRSA) can be thought of as one of the original “superbugs”, but despite marked falls in invasive infections in some settings, including the United Kingdom (UK) (Duerden et al., 2015), it remains a serious threat.

Efforts to control the spread and impact of AMR depend on understanding the transmission of resistant pathogens and the impact antimicrobial use. The relationship between human antimicrobial use and population-level risk of AMR is well established, e.g. (Seppälä et al., 1997). Similarly increased individual use of antimicrobials is associated with increased personal risk of AMR including, e.g., AMR in nasally carried *S. aureus* (van Bijnen et al., 2015). Time series approaches have been used to show temporal relationships between several antimicrobial classes and MRSA incidence (Vernaz et al., 2008, Aldeyab et al., 2008). However, to date the specific impact of antimicrobial exposures on individual patient transmission dynamics within healthcare settings has not been explored.

Mathematical modelling can provide powerful tools for understanding the transmission of healthcare-associated infections and other drug-resistant pathogens, and in a number of cases has directly informed national control policies ((Robotham et al., 2011), (van Bunnik et al., 2014), (Bootsma et al., 2011)), but wider application of models to inform AMR control polices requires an improved biological understanding of the key epidemiological and evolutionary processes (Knight et al., 2019). Amongst the most important needs is for a quantitative understanding for how patient antimicrobial exposure selects for resistant organisms. Mechanistically this involves understanding at an individual patient level the extent to which antimicrobials affect susceptibility to infection and whether they promote or inhibit onward transmission. Antimicrobials may also change detection of pathogens in screening or clinical testing. If this is not accounted for it can lead to erroneous conclusions, for example, pathogen colonisation temporarily supressed at hospital admission by antimicrobials may falsely be attributed to healthcare-associated transmission if it becomes apparent later in a hospital admission when antimicrobials are stopped (Price et al., 2017).

Temporal trends in healthcare associated infections are typically reported in terms of changes in incidence, rather than analysing underling transmission processes. The marked fall in MRSA incidence in the UK followed the introduction of a bundle of intensive control interventions and mandatory reporting of MRSA blood stream infections, and it is plausible that these measures contributed to the decline of MRSA (Duerden et al., 2015). However, it is also possible that at least part of the decline may have occurred in the absence of such interventions, perhaps driven by long-term ecological interactions between components of the nasopharyngeal flora (Wyllie et al., 2011). Mathematical modelling offers the opportunity to study how specific processes have changed over time, for example how infectious a given patient on a hospital ward is. Better understanding the relative contribution of decreased transmission in hospitals versus decreased MRSA importation from the community may yield insights into the relative contribution of hospital infection control or ecological changes.

Here we present a statistical inference approach that allows transmission events to be reconstructed and the impact of antimicrobial exposures on acquisition, onward transmission and detection to be estimated. Our approach also accounts for changes over time in transmission and importation. We use a data-augmented Markov chain Monte Carlo (MCMC) method to allow for the fact we do not directly observe transmission, but instead a series of imperfectly sensitive screening results, alongside patients’ hospital admission records. We apply our approach to study the transmission of MRSA in an adult intensive care unit (ICU) in Oxford, UK, between 2008 and 2017, allowing insights to be gained into the mechanisms behind the successful control of MRSA in the UK during this period. We also extend our models to allow discrete subtypes of MRSA to be modelled and show how antimicrobial susceptibility data can be used to inform transmission estimates.

## Results

We model the transmission of MRSA within a hospital intensive care unit (ICU), regarding patients as either colonised (with probability φ) or susceptible at the point of admission to the ICU. Once admitted susceptible patients can become infected at a rate proportional to the number of colonised patients present in the ICU (*βC*). Patients are screened for MRSA at admission and then at regular intervals; the screening test is assumed to be imperfectly sensitive (with sensitivity *ρ*), i.e. false negative results can occur, but perfectly specific, i.e. there are no false positive results. The analysis is performed in discrete time using daily time steps reflecting the precision of some of the underlying data items. Exposure to antibiotics alters the rate of onward transmission from each colonised patient by a factor, *τ*, susceptibility to colonisation in susceptible patients by a factor, *α*, and test sensitivity by a factor, *δ*.

### Identifying the effect of antibiotics

We use simulated data to show that given a large enough dataset our approach was able to independently and simultaneously identify all three effects of antibiotics specified above (Figure 1). We generated 10 datasets with 10000 admissions during 2000 days and assumed the true effect of antibiotics on onward transmission, acquisition and detection to be relatively modest, i.e. *τ* =1.2, *α* =1.3 and *δ* =1.1 respectively. The circulating MRSA was assumed to be from one of four types present at equal frequency, and model inferences were generated with and without accounting for this typing data. The models were also able to successfully recover the main model parameters (φ,*β,ρ*) (Supplementary Figure 1) and the statuses of the patients (Supplementary Figure 2). We also generated 40 datasets with time dependent transmission and importation and were able to recover increases as well as decreases in both transmission and importation (Supplementary Figure 3). Accounting for typing improves estimates of the importation parameter (Supplementary Figure 1), but does not yield more precise estimates of the antibiotic effects (Figure 1). Estimates of the effect of antibiotics on detection had the narrowest credibility intervals, reflecting the fact that data from all patients with positive swabs, who may be tested multiple times, are informative here. There was also less uncertainty about the impact of antibiotics on acquisition than onward transmission, likely reflecting the increased uncertainty introduced by the multiple possible sources for many acquisition events.

**Figure 1.**
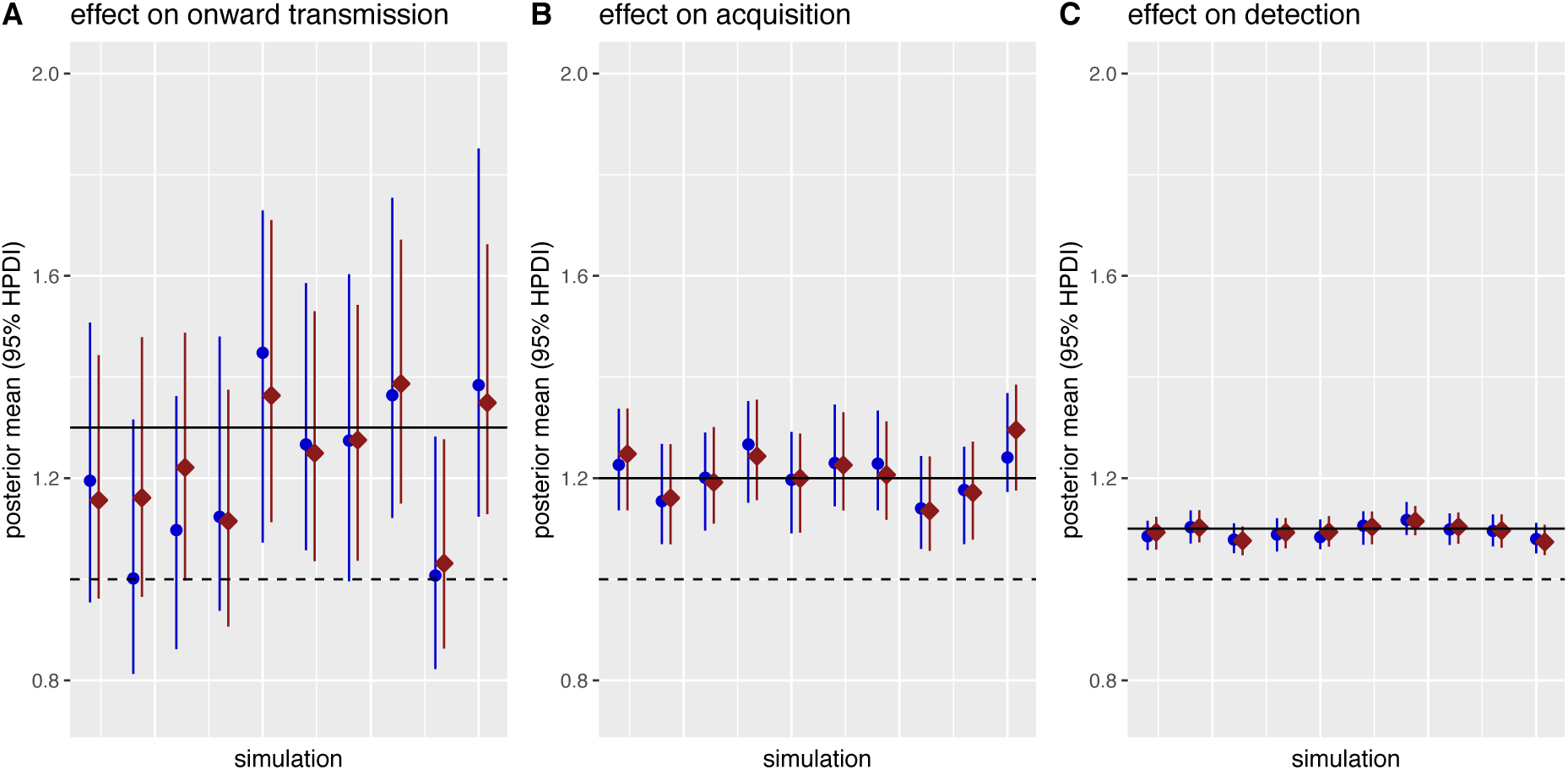
Posterior estimates of the effect of antibiotics on onward transmission, acquisition and detection in 10 simulated datasets. The true values are indicated by the solid horizontal lines. Each dataset was analysed with the model using positive and negative swabs only (blue circles) and using typing information (red squares, assuming 4 types were present at equal frequency). The dashed horizontal lines indicate 1 (i.e. no effect). HPDI, highest posterior density interval.

**Figure 2.**
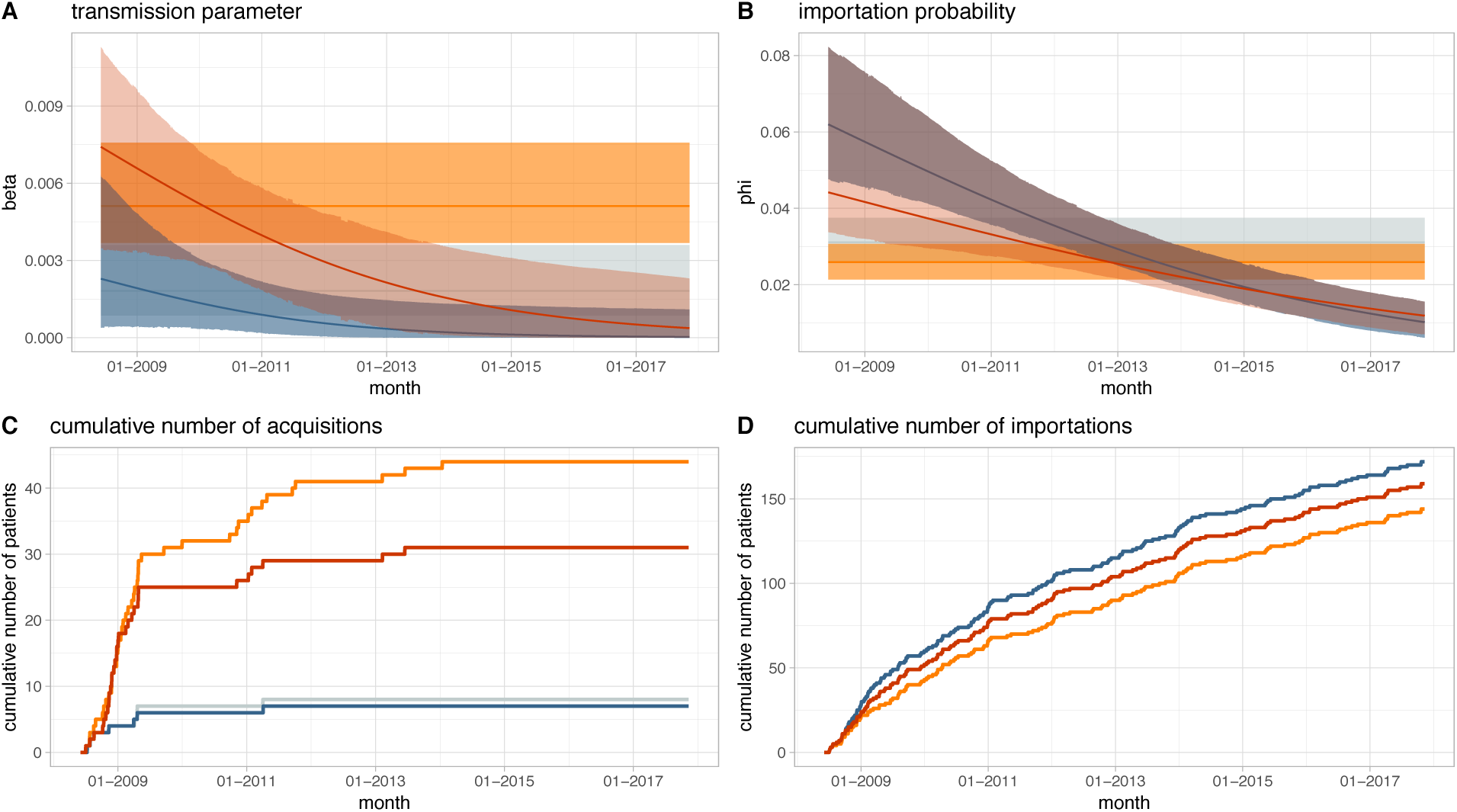
Change in transmission and importation of MRSA in an Oxford ICU, 2008-2017: posterior estimates from four different models. The top panels show point estimates (solid lines) and highest posterior density intervals (ribbons) of the transmission (A) and importation (B) parameters from the models with constant transmission and importation without (grey) and with (orange) considering antibiograms as typing data, and the models with time dependent transmission and importation without (blue) and with (red) antibiograms. The bottom panels show the posterior mean estimates of the number of patients estimated to have been colonised due to acquisition (C) and importation (D). The models without typing data (grey and blue) estimated very similar numbers of acquisitions and importations.

**Figure 3.**
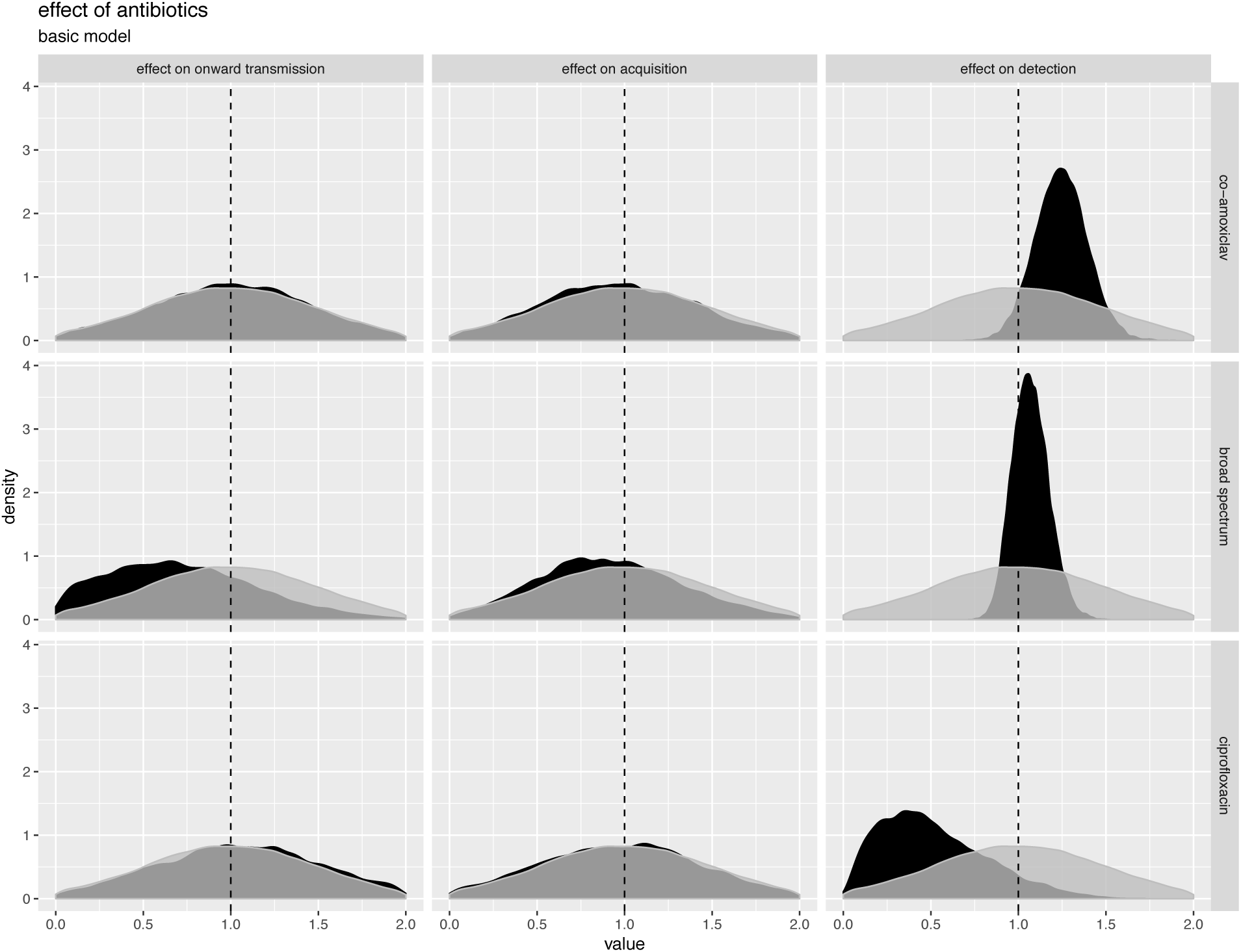
Effects of antibiotics on acquisition, onward transmission and MRSA detection in an Oxford ICU, 2008-2017. Posterior estimates are shown in black and prior distributions in grey. The plot is based on a model without time-dependent transmission or importation parameters or typing data. Alternative plots for these models are very similar and shown in Supplementary Figures 6-8.

We also analysed simulations including re-admissions of a subset of the same patients to the ICU. In the baseline model described above these re-admissions were artificially treated as new patients. This simplifying assumption avoided the need to track the colonisation status of patients between ICU admissions. We also developed an alternative formulation that tracked the probability of previously admitted patients being re-admitted colonised, described in Supplementary File 1. Model inferences were similar for both models (Supplementary File 1, Figure i), and as the baseline model was computationally simpler this was used for subsequent analyses. The proportion of patients readmitted to the ICU in our study dataset was relatively low (around 10%), using this proportion in simulations produced similar parameter estimates using both approaches, however in settings with higher re-admission rates the more complex model may be required.

### Analysing MRSA transmission and antibiotic effects in Oxfordshire, UK

We analysed data from the Oxford University Hospitals NHS Foundation Trust, a 1200-bed teaching hospital group providing secondary care to the population of Oxfordshire, UK (approximately 600000 people), and tertiary care services to the surrounding region. Data were available for all patients admitted to the combined medical and surgical adult ICU between June 2008 and November 2017 inclusive. Data items included ICU admission and discharge dates, MRSA screening swab results including antimicrobial susceptibility testing results for positive cultures, antimicrobial prescriptions, and patient factors including age, sex, Charlson comorbidity score, and admission specialty (Supplementary Table 1).

**Table 1.** Model parameters and priors.

| parameter | interpretation | value in simulations | prior |
| --- | --- | --- | --- |
| $\beta$ | transmission parameter | 0.02 | Half-Cauchy (0, 4) |
| $\rho$ | test sensitivity | 0.8 | Uniform (0, 1) |
| $\varphi$ | proportion of patients admitted already colonised (community prevalence) | 0.1 | Beta (Gibbs sampling) |
| $\alpha$ | effect of antibiotics on acquisition | 1.3 | Normal (1, 0.5) |
| $\tau$ | effect of antibiotics onward transmission | 1.2 | Normal (1, 0.5) |
| $\delta$ | effect of antibiotics on detection | 1.1 | Normal (1, 0.5) |
| $a_\varphi$ | upper asymptote of importation in time dependent model | 0.2 | Half-Normal (0, 0.1) |
| $b_\varphi$ | slope of importation in time dependent model | -0.05 | Normal (0, 0.001) |
| $c_\varphi$ | horizontal shift of importation in time dependent model | 1 | Normal (0, 100) |
| $a_\beta$ | upper asymptote of transmission parameter in time dependent model | 0.1 | Half-Normal (0, 0.1) |
| $b_\beta$ | slope of transmission parameter in time dependent model | -0.02 | Normal (0, 0.001) |
| $c_\beta$ | horizontal shift of transmission parameter in time dependent model | 1 | Normal (0, 100) |

There were 7924 admissions to the ICU from 7138 patients. A total of 12047 MRSA screens were performed in 6757 patients, 271 (2.2%) were MRSA-positive in 179 different patients. An overview of the antimicrobial prescription data is shown in Supplementary Figure 4, the most commonly prescribed antibiotics were co-amoxiclav, vancomycin, metronidazole, piperacillin-tazobactam and meropenem. Molecular typing or whole-genome sequencing was not routinely undertaken, however routine antimicrobial susceptibility testing provides a proxy for isolate relatedness. In addition to confirming methicillin resistance, seven antibiotics (gentamicin, erythromycin, tetracycline, fusidic acid, ciprofloxacin, rifampicin and mupirocin) were consistently tested for during the whole study period and therefore used to establish resistance profiles. The number of tests per resistance profile and the distance between profiles over time for individual patients are shown in Supplementary Figure 5. We found 31 different combinations of presence and absence of resistance with respect to these seven antibiotics. The maximum number of profiles in a single patient was 5. However, no patient had multiple different profiles during the same admission. Therefore, as repeat admissions were considered separately, when using typing data, we considered transmission from one patient to another to be plausible only when both patients were colonized with strains with identical resistance profiles. We also performed analyses without typing data, which provides a sensitivity analysis if the assumption requiring matching antibiograms is overly restrictive. For 50 admissions with a positive test there was no profile as recorded susceptibly data was not present for all seven antibiotics. These profiles were augmented in the same way that profiles for patients with no positive test were augmented, which is described in more detail in the Methods section.

**Figure 4.**
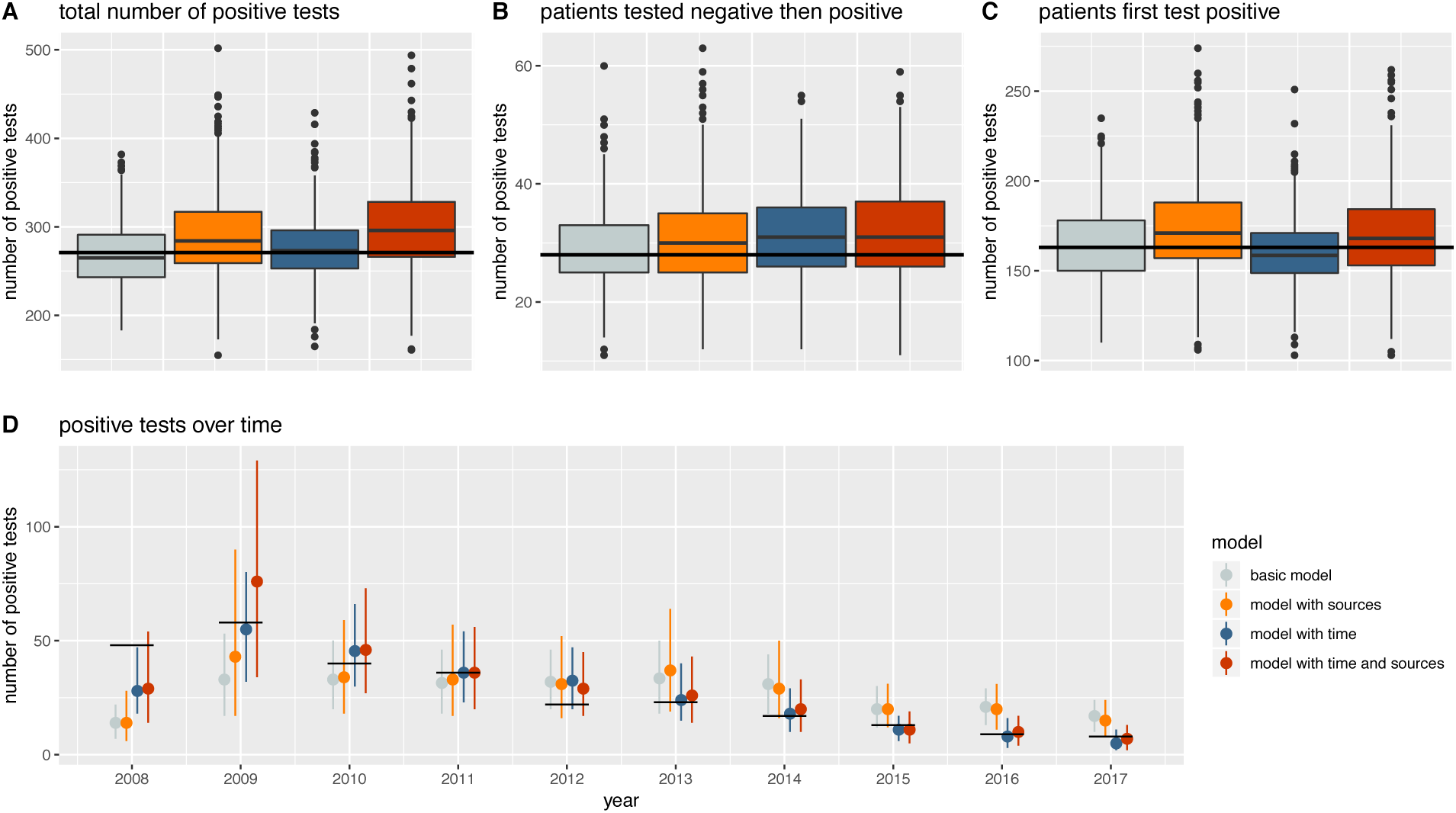
Posterior predictive checks. The number of positive tests in the true patient data (black lines) is compared to the positive tests in 1000 simulated datasets (boxplots, mean and interquartile range). D shows the number of positive tests for each year in the true patient data (black lines) and in 1000 simulated datasets (median and 0.9 credible interval).

#### Time dependent transmission and importation

The number of positive tests per year decreased over time from 48 in 2009 to 9 in 2016, while the diversity of the resistance profiles observed increased with time (Supplementary Figure 6). While in the earlier years of the study, a majority of colonized patients had one of two resistance profiles (ciprofloxacin resistance only or ciprofloxacin and erythromycin resistance) all 8 colonized patients in 2017 had a different resistance profile. Provided transmission events associated with changes in antibiograms are uncommon, as suggested by the stability of antibiograms over time in individual patients (Supplementary Figure 5), the diversity of resistance profiles seen at the end of the study suggests that towards the end of the study transmission was uncommon and importation was most importation driver of positive tests. We formally model this by allowing the transmission and importation parameters to vary with time. We find a strong decrease in both the transmission parameter and the importation probability over time (Figure 2). In the model using typing data, the estimate of *β* falls from 0.0074 (95% highest posterior density interval, HPDI 0.0035, 0.0113) in 2008 to 0.0004 (1.9e-7, 0.0023) by 2017 (Figure 2A). This is reflected in the number of acquisitions from early 2013 onwards being estimated as zero (Figure 2C). The importation probability falls from 0.044 (0.034, 0.060) in 2008 to 0.012 (0.007, 0.018) in 2017 (Figure 2B and 2D). The signal for these declines is captured by the slope of the function we use to model time, which is −0.0014 (−0.0032, −0.0002) and −0.0007 (−0.0009, − 0.0005) for transmission and importation respectively (Supplementary Figure 7). Temporal trends in transmission and importation from the model without typing data were similar (Figure 2, which also shows estimates from models without time-dependent transmission and importation).

#### Inferring effects of antibiotics

We analysed the effects of antibiotics for 3 antibiotics/antibiotic groups: co-amoxiclav (the most commonly used), ciprofloxacin, and broad-spectrum beta-lactams without activity against MRSA (including co-amoxiclav, piperacillin/tazobactam, cefuroxime, ceftriaxone, eftazidime, ceftolozane and meropenem). As a simplifying assumption, the effects were considered to last only while the patient was taking the antibiotics. The posterior estimates for the effect of antibiotics on transmission, acquisition and detection are shown in Figure 3, using the basic model without time-dependent transmission or typing data. (The very similar posterior estimates from the model with typing data and the model with time dependent transmission and importation are shown in Supplementary Figures 8-10.)

There was a moderate effect of antibiotics on detection. Co-amoxiclav was associated with a 1.3-fold (95% HPDI 1.01, 1.57) increase in the relative probability of detection, however there was no strong evidence that this was also the case for broad-spectrum beta-lactams as a group,1.03-fold (95% HPDI 0.87, 1.26). The proportion of patients who were given an MRSA active drug simultaneously with any broad-spectrum beta-lactam antibiotic or co-amoxiclav on the day of the test was 0.25 and 0.25 respectively. Conversely, ciprofloxacin was estimated to be associated with a 0.29-fold (95% HPDI 0.03, 1.04) reduction in test sensitivity. However only a handful of tests were conducted while patients were exposed to ciprofloxacin, one positive test and an estimated four false negative tests, and the uncertainty was large. The true positive test was a result of colonisation with ciprofloxacin-resistant MRSA, and the inferred resistance profiles of the four false negative results also included ciprofloxacin resistance, so a reduction in sensitivity due to ciprofloxacin being active against MRSA is unlikely.

Posterior distributions of the effects of antibiotics on acquisition or onward transmission of MRSA were very similar to the priors, and for all three groups of antibiotics considered we are unable to rule out substantial effects in both directions (Figure 3). For example, co-amoxiclav was estimated to be associated with a 0.88-fold reduction in acquisition (95% HPDI 0.22, 1.66) and a 1.20-fold increase in onward transmission (95% HPDI 0.22, 1.87) respectively. This lack of information about the effects of antibiotics on acquisition and transmission rates reflects the low number of possible transmission events in the data. Of the 179 patients who were tested positive, only 50 were not tested positive on admission, which is many fewer than the transmission events used to identify the antibiotic effects in simulated data.

#### Patient factors associated with acquisition and onward transmission

We used the status (susceptible, imported, acquired) with the highest posterior density of each patient and performed logistic regression to investigate for any relationship between importation, acquisition or onward transmission and patient age, sex, Charlson comorbidity score, admission speciality group (acute medicine, specialist medicine, general surgery, trauma and orthopaedics [T&O], other). Controlling for all other factors, patients admitted under T&O were less likely to be admitted colonised than the acute medicine reference group (OR 0.52 [95%CI 0.28, 0.97]) and importation was associated with increased age (OR per 10 year increase 1.11 [1.01, 1.22]). There was no strong statistical evidence that any of the other variables studied were associated with a change in importation, acquisition and onward transmission. The results for all variables studied are reported in Supplementary Table 2.

**Table 2.** Absolute difference between the maximum posterior density point estimate and the true value (top row, median and interquartile range) from ten simulated datasets and proportion of simulations where the true value lies within the 0.90 highest posterior density interval (bottom row). The parameters a,b and c in for the time dependent models refer to the scaling parameters of the time dependent transmission and importation function, as described in the methods section.

| | transmission parameter ($\beta$) | importation probability ($\varphi$) | effect on acquisition ($\alpha$) | effect on transmission ($\tau$) | test sensitivity ($\rho$) | effect on detection ($\delta$) |
| --- | --- | --- | --- | --- | --- | --- |
| **basic model** | 0.0034 [0.0009, 0.0042]<br>1 | 0.0147 [0.0093, 0.0209]<br>1 | 0.1405 [0.0940, 0.3120]<br>0.9 | 0.1317 [0.0554, 0.2441]<br>1 | 0.0700 [0.0137, 0.0872]<br>0.9 | 0.0851 [0.0460, 0.0942]<br>1 |
| **model with sources** | 0.0017 [0.0006, 0.0032]<br>1 | 0.0117 [0.0109, 0.0156]<br>1 | 0.1729 [0.0595, 0.3858]<br>1 | 0.2027 [0.1204, 0.2532]<br>1 | 0.0566 [0.0344, 0.0806]<br>0.9 | 0.0841 [0.0445, 0.0902]<br>1 |
| **model with time** | $a_\beta$<br>0.0115 [0.0049, 0.0132]<br>0.9<br>$b_\beta$<br>0.0009 [0.0007, 0.0011]<br>1 | $a_\varphi$<br>0.0357 [0.0152, 0.0535]<br>0.9<br>$b_\varphi$<br>0.0010 [0.0009, 0.0011]<br>1 | 0.1482 [0.0500, 0.2238]<br>1 | 0.2540 [0.0913, 0.4509]<br>1 | 0.0597 [0.0196, 0.0895]<br>0.9 | 0.0581 [0.0272, 0.0827]<br>1 |
| | 1<br>$c_\beta$<br>12.2 [5.9, 20.2]<br>1 | $c_\phi$<br>11.1 [4.1, 17.9]<br>1 | | | | |
| **model with time and sources** | $a_\beta$<br>0.0100 [0.0058, 0.0147]<br>0.9<br>$b_\beta$<br>0.0009 [0.0007, 0.0011]<br>1<br>$c_\beta$<br>9.2 [3.0, 12.2]<br>1 | $a_\phi$<br>0.0178 [0.0029, 0.0297]<br>1<br>$b_\phi$<br>0.0009 [0.0007, 0.0011]<br>1<br>$c_\phi$<br>13.7 [5.9, 18.1]<br>1 | 0.1483 [0.0909, 0.2203]<br>1 | 0.2778 [0.1342, 0.5164]<br>1 | 0.0545 [0.0268, 0.0702]<br>0.9 | 0.0600 [0.0301, 0.0845]<br>1 |

## Discussion

We describe an individual-patient probabilistic method for simultaneously reconstructing transmission events, estimating parameters and quantifying important co-variates. We apply this to study healthcare-associated MRSA transmission, the impact of antibiotics and changes over time.

We make several key observations. Firstly, it is possible to independently recover the effects of antibiotics on acquisition, onward transmission and detection, even if these changes are relatively modest. However, this requires large datasets e.g. in our simulations several thousand MRSA cases were needed, of which up to 80% were acquisitions which facilitated estimation of acquisition/onward transmission effects. In many settings this is likely to exceed the number of cases present within a dataset, unless potential heterogeneity is introduced by pooling data across multiple institutions or over prolonged periods of time (which would need to be accounted for). In the dataset we study, there are an order of magnitude fewer cases. This suggests that to make reliable estimates of the effect of antibiotics on transmission we are likely to need to pool data from multiple wards or hospitals, taking care to appropriately to account for heterogeneity. Here, we can only rule out very large effects, e.g. increases in acquisition or onward transmission of >1.9-fold and >1.7-fold respectively.

We find evidence that antibiotics can have contrasting associations on MRSA detection, with co-amoxiclav associated with enhanced detection and ciprofloxacin associated with reducing detection. These findings may reflect co-amoxiclav, without activity against MRSA, clearing other organisms present in the nose and allowing MRSA to proliferate. However, why the same effect was not seen with all broad-spectrum beta-lactams is unclear. Additionally, with much of the MRSA isolated resistant to ciprofloxacin it seems unlikely that the ciprofloxacin effect is causal unless *in vitro* test results are not reflecting *in vivo* activity. Antibiotic exposure transiently masking pre-existing carriage has been hypothesized to account for some apparent hospital acquisition on the basis of whole genome sequencing (Price et al., 2017), and these findings underscore the need to better understand the role of antibiotics on detection in transmission studies.

We find that importation of MRSA decreased from around 5% in 2008 to below 2% in 2018. This is consistent with estimates of importation found in other studies. Forrester et al., 2007 found an importation probability of 4.6% in an ICU in London between 1995 and 1997 and Worby et al., 2016 estimated the importation probability in a neonatal ward in Cambridge at around 1.2% in 2011.

We explore the relative contribution of falls in transmission and importation to the decline of MRSA seen on our ICU, mirroring the decline in MRSA in the UK nationally. We find evidence of transmission rates falling from 2009 onwards. In fact, it is likely that transmission rates had been falling prior to our study, given the timing of key interventions locally and nationally earlier in the decade (Duerden et al., 2015). In the ICU studied, most infection control enhancements made in response to MRSA were already in place at the start of the study, including isolation and contact precautions for colonised patients, screening of all patients at ICU admission and at regular intervals, universal skin decolonisation with 2% chlorhexidine, antimicrobial impregnated central lines, with dedication insertion packs and care standards and root-cause analyses of all blood stream infections. Further details of these interventions can be found in (Wyllie et al., 2011).

Our study has several limitations in addition to the limited power to detect antibiotic effects. Rather than model re-admissions, as in our setting these are uncommon to the ICU, we treat re-admissions as new patients. However, we have detailed an alternative approach to account for these, but it is more computationally intensive. Our model also treats the ICU in isolation to the rest of the hospital, as antimicrobial prescribing data were only available for the ICU. However, MRSA transmission dynamics within the hospital as a whole likely contributed to MRSA observed on the ICU. We use antibiograms as an example of a discrete typing scheme, however this is a relatively crude typing scheme, that may vary within a single infection (Eyre et al., 2012), although we did not see this in the current study. However, our approach could be extended to other typing schemes, such as *spa*-typing or multilocus sequence typing. We also did not study the effect of glycopeptides including vancomycin and teicoplanin directly. This was a prospectively made decision given concern about reverse causation whereby patients suspected (or known, from another hospital’s data) to have MRSA colonisation may be given these antibiotics prior to a positive test, which would complicate the interpretation of the observed increase in test sensitivity associated with vancomycin administration (Supplementary Figure 12).

We use data which is routinely collected in an ICU. This has the advantage that our model could potentially be applied to other, similar datasets. But as with all such observational studies we cannot make strong causal statements about antibiotics effects as they were not randomly assigned to patients. Our modelling approach expands existing methods by allowing to estimate the effect of covariates. This could be further developed by capturing effects other than multiplicative and by extending the timespan of effects beyond the day of antibiotic exposure.

In conclusion, we present a method that can allow important risk factors for transmission and infection detection to be estimated. This provides a mechanism for undertaking data-driven infection control whereby not just who-infected-whom can be estimated, but also the conditions leading to transmission understood and combatted. This is likely to lead to better infection control in future and in turn better outcomes for patients.

## Methods

### Transmission models and likelihood functions

We implement a stochastic mechanistic transmission model, that builds on previous approaches, e.g. (Forrester et al., 2007, Worby et al., 2016). The data augmentation approach used in these publications introduces additional parameters for the unknown status of for each patient, which makes it possible to split the overall likelihood into the following product:

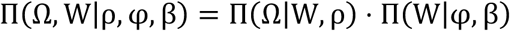

where Ω denotes the test results, W is the colonisation status of the patients and colonisation times, *ρ* is the test sensitivity, *φ* is the probability of a patient being positive on admission and *β* is a transmission parameter. The first factor of the product describes the observation process, which can be modelled as a binomial distribution. Assuming perfect specificity, the likelihood of the test results given the test sensitivity and the true statuses is therefore given by

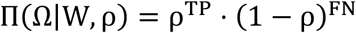

where TP and FN denote the number of true positive and false negative tests. The second factor of the overall likelihood captures the transmission process. Following (Worby et al., 2016) we assume that patients who acquire a colonisation stay colonised until discharge. The patient statuses, W, are therefore fully described by the colonisation times, *t*^*c*^, which are set to infinity for patients who do not get colonised during their ward stay. By discretising time into days, the likelihood of the colonisation times given the transmission parameters can be modelled as

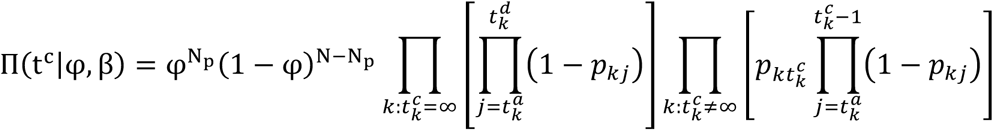

where N is the total number of patients, N_p_ is the number of patients who are admitted already colonised and 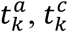 and 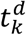 denote the day of admission to the ICU, colonisation and discharge from the ICU of patient k. *p*_*kj*_is the probability of patient k becoming colonised on day j which is given by

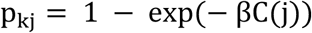

where C(j) is the number of colonised patients present on the ICU on day j.

### Accounting for the effect of antibiotics

We expand this model to account for the effect of antibiotics on detection and transmission. We assume that antibiotics can affect the test sensitivity, the probability of acquisition and the probability of onward transmission. To capture the effect of antibiotics on test sensitivity we assume that in the presence of antibiotics the baseline test sensitivity changes by a factor *δ* and therefore model the likelihood of the test results as

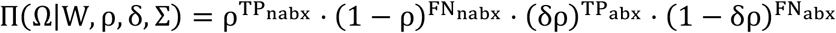

where *TP*_*nabx*_, *FN*_*nabx*_, *TP*_*abx*_and *FN*_*abx*_denote the total number of true positive and false negative tests conducted while a patient was off or on antibiotics and Σ is a matrix with entries σ_*ij*_ equal to one if patient i is on antibiotics on day j and zero otherwise. We model the effect of antibiotics on the transmission dynamics by introducing a parameter *α* for the effect on acquisition and a parameter *τ* for the effect on onward transmission and modify the daily probability of acquisition as follows

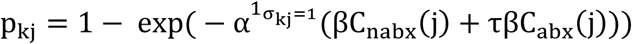

where *C*_*nabx*_(j) and *C*_*abx*_(j) denote the number of colonised patients on day j who are off or on antibiotics and 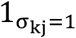 is an indicator function which equals to 1 if patient k is on antibiotics on day j and zero otherwise.

### Modifications to the likelihood and data augmentation with typing data

To extend our model to account for discrete typing data we use a simplified version of the model by (Worby et al., 2016)). If the testing contains some additional information on the type of infection this information can be used to determine likely sources of infection and therefore potentially increase the overall precision of the estimates. The dataset we are using does not contain typing or sequencing data, however, resistance profiles can be used as a proxy for the type of strain. We denote by Z the augmented source patients for all transmission events and factorise the overall likelihood as follows

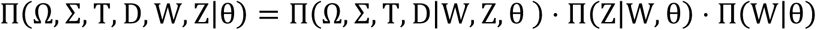

with T denoting the MRSA type causing colonisation and θa vector whose elements are the model parameters β,ρ,φ,α,δandτ described in Table 1. D is a matrix of distances between the different types. In the simplest case we can assume that patients colonised with different MRSA types cannot be linked by transmission and set the likelihood contribution of all patients who acquire to 1 if the augmented type of the patient is the same as the type as the source and to 0 otherwise. For patients who are imported and do not have a positive test, a type has to be augmented. We assume that for the imported patients the probability of being colonised with any given type is equal to the proportion of patients who were tested positive and found to have that type. We therefore model the likelihood of the observed types given the augmented sources and colonisation times as

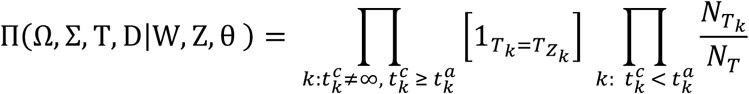

where 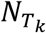 is the number of patients who were tested positive with type *T*_*k*_and *N*_*T*_is the total number of patients tested positive with any type. The first term applies to patients who acquire the pathogen in the ward. Since we assume that transmission can only take place between patients with the same type, this term, denoted by the indicator function 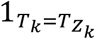, is zero if the type of any patient, *T*_*k*_, is not equal to the type of the source of infection for that patient, 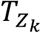.The second term is the importation model. Since our model allows for an effect of antibiotics on onward transmission, patients who are on antibiotics on a given day are not equally likely to act as sources of infection as patients who are not on antibiotics. In the absence of effects of antibiotics, the likelihood of any given patient being the source of infection to a patient who acquires infection on a given day is equal to one over the number of colonised patients on that day. In that case the likelihood of the sources given the colonisation times would be given by

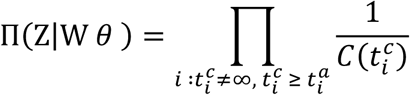

where 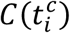 is the number of colonised patients on the day when patient i becomes colonised. To account for the effect of antibiotics on transmission we modify this as follows

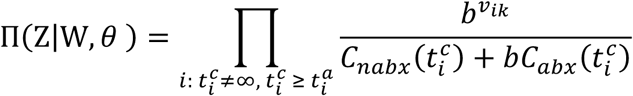

with

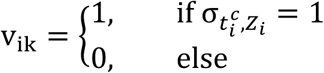

The approach we present here can also be used for discrete typing schemes such as multi-locus sequence typing (MLST). It can also be easily adapted to more continuous measures of distance, such as *spa* typing, by deriving a likelihood function which is based on the distance between the type of the offspring and the type of the proposed source.

### Time dependent acquisition and importation

We are analysing a dataset that was collected during 10 years, a timespan during which changes in importation and transmission probability are likely to occur. We can account for this by allowing the importation probability, φ, and the transmission parameter, β, to vary with time and set

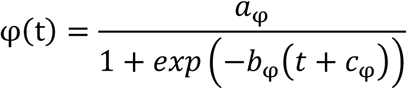

and

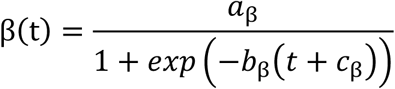

with parameters *a*_*φ*_,*b*_*φ*_,*c*_*φ*_,*a*_*β*_,*b*_*β*_ and *c*_*β*_ being estimated. An example of curves resulting from different combinations of the shape parameters a, b and c is shown in Supplementary Figure 11.

#### MCMC algorithm and implementation

We use a Markov chain Monte Carlo (MCMC) algorithm to generate posterior estimates of the model parameters and the augmented patient statuses. In each iteration of the MCMC we first update the parameters of the transmission model one at a time using a Metropolis step and the importation probability via a Gibbs update. We then update the statuses and sources of infection of a subset of patients. The moves and Hastings ratios involved in the status updates are described in detail in Worby et al., 2016. The width of the proposal distributions are adapted during the first 30% of the iterations in order to achieve an acceptance ratio between 0.1 and 0.3 for each parameter. These iterations are discarded as part of the burn-in. Convergence is assessed using the Gelman-Rubin convergence diagnostic as implemented in the Rpackage coda using 1.1 as a cut-off indicating convergence. We combine chains from different starting values and use a minimum effective sample size of above 200 for all parameters. We use weakly informative priors for all parameters. Parameters and priors are shown in **Table 1**.

The models are implemented in R and C++, making use of Rcpp (Eddelbuettel and François, 2011). All four models are available as a package (https://github.com/mirjamlaager/mrsamcmc). The package also includes functions for forward simulations which can be used to create simulated data, conduct posterior predictive checks and simulate the effect of interventions.

#### Simulations

We use simulated data to show that our models are accurate and well calibrated. For each model we generate 10 datasets, simulating under the same assumptions as in the inference model. For each dataset we compute the absolute difference between the point estimate and the true value of the parameter (precision) and check whether the true value lies within the 90% highest posterior density interval (calibration). The average precision and calibration are shown in Table 2.

#### Oxford ICU dataset

Data were extracted from an anonymised extract of linked admissions to and microbiology data from the Oxford University Hospitals NHS Foundation Trust from the Infections in Oxfordshire Research Database (IORD) which has generic Research Ethics Committee, Health Research Authority and Confidentiality Advisory Group approvals (14/SC/1069, ECC5-017(A)/2009).

#### Model Assessment

We assessed the model fit in our Oxford ICU analysis by sampling 1000 parameters from the posterior distributions and running forward simulations with the simulated values. These posterior predictive checks have been advocated for as a useful tool to assess whether a model is able to reproduce key aspects of the transmission process (Gibson et al., 2018). The results are shown in Figure 4. The total number of positive tests in the dataset lies within the interquartile range of the simulated data for all four models. To check whether the models appropriately differentiate between acquisition and importation we used the number of patients with a negative test followed by a positive test as an approximation of acquisitions and the number of patients whose first test was positive as an approximation of importations. All four models performed well in this comparison. The decrease of positive tests over time is better captured by the models that include time.

## Data Availability

Data on patient hospital admissions cannot be shared publicly under the terms of the approvals granted for their use. Applications for access to these data can be made via the Infections in Oxfordshire Research Database https://oxfordbrc.nihr.ac.uk/research-themes-overview/antimicrobial-resistance-and-modernising-microbiology/infections-in-oxfordshire-research-database-iord/ by researchers who meet the criteria for access to confidential data.

## Acknowledgements

We thank all the people of Oxfordshire who contribute to the Infections in Oxfordshire Research Database. Research Database Team (Oxford): R Alstead, C Bunch, DW Crook, J Davies, J Finney, J Gearing (community), H Jones, L O’Connor, TEA Peto (PI), TP Quan, J Robinson (community), B Shine, AS Walker, D Waller, D Wyllie. Patient and Public Panel: G Blower, C Mancey, P McLoughlin, B Nichols.

